# Neurobiological correlates of longitudinal grey matter volume changes in preclinical Alzheimer’s disease

**DOI:** 10.64898/2025.12.01.25341227

**Authors:** Wiesje Pelkmans, Raffaele Cacciaglia, Michalis Kassinopoulos, Oriol Grau-Rivera, Carolina Minguillón, José Luis Molinuevo, Gwendlyn Kollmorgen, Clara Quijano-Rubio, Rachel E. Wilson, Erin M. Jonaitis, Rebecca E. Langhough, Kaj Blennow, Henrik Zetterberg, Marc Suárez-Calvet, Gemma Salvadó, Sterling C. Johnson, Juan Domingo Gispert

**Affiliations:** Barcelonaβeta Brain Research Center (BBRC), Pasqual Maragall Foundation, Barcelona, Spain; Hospital del Mar Research Institute, Barcelona, Spain; Centro de Investigación Biomédica en Red de Fragilidad y Envejecimiento Saludable (CIBERFES), Instituto de Salud Carlos III, Madrid, Spain; H. Lundbeck A/S, Copenhagen, Denmark; Roche Diagnostics GmbH, Penzberg, Germany; Roche Diagnostics International Ltd, Rotkreuz, Switzerland; Wisconsin Alzheimer’s Disease Research Center, University of Wisconsin School of Medicine and Public Health, Madison, WI, USA; Department of Medicine, Division of Geriatrics and Gerontology, School of Medicine and Public Health, University of Wisconsin–Madison, Madison, WI, USA; Wisconsin Alzheimer’s Institute, University of Wisconsin School of Medicine and Public Health, Madison, WI, USA; Department of Psychiatry and Neurochemistry, Institute of Neuroscience and Physiology, The Sahlgrenska Academy, University of Gothenburg, Mölndal, Sweden; Clinical Neurochemistry Laboratory, Sahlgrenska University Hospital, Mölndal, Sweden; Paris Brain Institute, ICM, Pitié-Salpêtrière Hospital, Sorbonne University, Paris, France; Neurodegenerative Disorder Research Center, Institute on Aging and Brain Disorders, University of Science and Technology of China and First Afgiliated Hospital of USTC, Heifei, China; Hong Kong Center for Neurodegenerative Diseases, Hong Kong, China; Department of Neurodegenerative Disease, UCL Institute of Neurology, London, United Kingdom; Servei de Neurologia, Hospital del Mar, Barcelona, Spain; Clinical Memory Research Unit, Department of Clinical Sciences, Lund University, Malmö, Sweden; Universitat Pompeu Fabra, Barcelona, Spain; Spanish National Center for Cardiovascular Research (CNIC), Madrid, Spain; Centro de Investigación Biomédica en Red Bioingeniería, Biomateriales y Nanomedicina (CIBER-BBN), Instituto de Salud Carlos III, Madrid, Spain

**Author notes:** corresponding authors: Dr. Pelkmans Dr. Salvadó.

**Keywords:** Amyloid-β, Astrocytic reactivity, Biomarkers, Cerebrospinal fluid, Cognitive decline, Grey matter volume, Microglial reactivity, Non-negative matrix factorization (NMF), Preclinical Alzheimer’s disease, Tau, Voxel-based morphometry (VBM)

## Abstract

**Background:** Structural brain changes during the earliest asymptomatic stages of Alzheimer’s disease (AD) remain poorly understood. Previous research in preclinical AD shows heterogeneous findings, reporting both subtle neuronal loss and paradoxical increases in grey matter (GM) volume. This study applies an extensive cerebrospinal fluid (CSF) biomarker panel to better understand the biological processes underlying longitudinal GM changes in cognitively unimpaired (CU) adults, spanning the amyloid/tau (AT) continuum.

**Methods:** We analysed data from 627 CU individuals from three longitudinal cohorts (ALFA+, Wisconsin ADRC, WRAP), with repeated MRI (3.5±0.9 years) and baseline CSF biomarkers from the NeuroToolKit panel (Roche Diagnostics). Using non-negative matrix factorization, we decomposed the CSF biomarker levels into six latent components, reflecting amyloid-β (Aβ) pathology, tau-related pathophysiology with synaptic injury, neuroaxonal injury, microglial reactivity, astrocytic reactivity, and cytokine signalling. We tested associations between component weights and voxel-wise longitudinal GM volume changes using single-component and a joint-all components model. Analyses were performed across the full sample and stratified by AT status. Associations with longitudinal cognitive performance (PACC) were assessed using linear mixed-effects models.

**Results:** The Aβ pathology component was the strongest and most widespread predictor of longitudinal GM atrophy, predominantly in temporal and frontal regions, also when controlling for tau pathophysiology, neuroaxonal injury, or neuroinflammatory components. Higher Aβ pathology scores were also associated with cognitive decline. The component capturing tau-related pathophysiology and synaptic injury initially associated with GM loss but lost significance after accounting for other biomarker components. In contrast, components reflecting microglial reactivity, astrocytic reactivity, and cytokine signalling were associated with longitudinal GM volume increases, with effects varying by AT stage.

**Conclusions:** In this large longitudinal sample of asymptomatic individuals, Aβ pathology emerged as the primary contributor to early neurodegeneration and cognitive decline, beyond the effects of tau pathophysiology and neuroaxonal injury. While glial and inflammatory processes may underlie transient GM increases in preclinical AD. A better understanding of these dynamic relationships between structural brain changes and diverse biological pathways at the earliest stages of AD is crucial to inform the development of interventions before irreversible neurodegeneration occurs.

## Background

Alzheimer’s disease (AD) is a progressive neurodegenerative disorder pathologically characterised by the accumulation of amyloid-beta (Aβ) plaques and tau neurofibrillary tangles, associated with widespread neuronal loss (1,2). Structural magnetic resonance imaging (MRI) has showed that this atrophy follows a stereotypical pattern, particularly involving medial temporal and associative cortices, that closely associate with the distribution of tau pathology (3–5). However, the precise biological mechanisms that drive structural brain changes, especially in early asymptomatic stages, remain poorly understood.

Numerous studies have reported heterogenous findings in cognitively unimpaired (CU) Aβ-positive individuals, with lower grey matter (GM) volume cross-sectionally (6–8) and subtle atrophy over time (9–12), but also paradoxical higher GM volume (13–18) and longitudinal volume increases (19,20). However, the significance and interpretation of these volume increases remain unclear, as such may reflect resilience, inflammatory swelling, or compensation, but could also result from selection bias, *i.e.* resilient individuals may tolerate more pathology while remaining cognitively unimpaired. Therefore, longitudinal MRI studies are crucial to capture within-person trajectories of neurodegeneration and non-linear or compensatory processes in the preclinical stages of AD (21,22). A better understanding of these early structural changes is particularly relevant as the field moves toward earlier diagnosis and disease modifying therapies in preclinical stages, before significant atrophy, tau pathology, or symptoms.

Most prior research examined single biomarkers in isolation, predominantly Aβ and tau, in relation to GM changes (23–26). While it is less well understood how other biological processes such as synaptic dysfunction, neuroaxonal injury, and neuroinflammation contribute to neurodegeneration (27). Incorporating multiple biomarkers that reflect diverse biological processes may better capture the complexity and heterogeneity of AD pathophysiology.

In this study we investigated the relationship between a comprehensive set of key cerebrospinal fluid (CSF) biomarkers reflecting diverse biological processes underlying AD pathophysiology and longitudinal GM volume changes in a large sample of CU individuals. Given the close interrelations and overlapping associations among many biomarkers and GM changes, we identified latent biomarker profiles capturing microglial reactivity, cytokine signalling, astrocytic reactivity, Aβ pathology, tau-related pathophysiology and synaptic injury, and neuroaxonal injury using non-negative matrix factorization (NMF). We then related these components to longitudinal voxel-wise GM volume changes over time, stratified by Aβ and tau biomarker status, in a large multi-cohort of CU individuals.

## Methods

### Participants

This study included data from three longitudinal cohorts of CU participants: ALFA+, WRAP, and the Wisconsin ADRC cohort. The ALFA+ (*For ALzheimer’s and Families*; Clinicaltrials.gov Identifier: NCT01835717) is a longitudinal monocentric research cohort conducted at the Barcelonaβeta Brain Research Center. It includes CU individuals enriched for apolipoprotein E *(APOE) ε4* allele carriership and family history of AD. Detailed descriptions of the ALFA parent cohort and inclusion criteria are available in Molinuevo et al. (2016). The Wisconsin Registry for Alzheimer’s Prevention (WRAP) and the Wisconsin Alzheimer’s Disease Research Center (ADRC) are longitudinal observational cohorts of participants who enrolled at midlife and are enriched with risk for late-onset AD due to parental history of AD dementia. These include individuals at various AD stages, from CU to AD dementia, and undergo annual or biennial detailed study evaluations. For a detailed description of the study designs and methods see (29,30). From the above-described cohorts, we selected individuals with unimpaired cognition (CDR=0) at baseline, available CSF biomarkers at baseline, and repeated volumetric MRI. Ethical approval was given by the regional ethics committees, and all research was completed in accordance with the Declaration of Helsinki.

### CSF biomarker measurements

CSF collection and processing were standardised across the ALFA+, WRAP, and Wisconsin ADRC cohorts. Lumbar puncture procedures followed standard guidelines (31), and all analyses were conducted at the Clinical Neurochemistry Laboratory, University of Gothenburg, Sweden. CSF core AD biomarkers (β-amyloid_40_, β-amyloid_42_, phospho-tau_181_ [p-tau]), markers of synaptic injury and neurodegeneration (neurogranin, neurofilament light protein [NfL], total-tau [t-tau], and α-synuclein), glial reactivity markers (glial fibrillary acidic protein [GFAP], chitinase-3-like protein 1 [YKL-40], soluble triggering receptor expressed on myeloid cells 2 [sTREM2], and S100 calcium binding protein B [S100B]), and inflammation markers (interleukin-6 [IL6]) were collected at baseline and quantified using the NeuroToolKit (NTK) (32), a panel of robust prototype assays (Roche Diagnostics International Ltd, Rotkreuz, Switzerland).

Participants were classified according to Aβ pathology (A; CSF Aβ_42/40_ ratio), and tau pathology (T; CSF p-tau_181_) status, based on predefined cut-offs (32,33). Individuals with non-AD pathological changes (*i.e.* A-T+, n=25) were excluded from further analysis.

The NTK project aims to generate robust and comparable biomarker data across multiple independent cohorts using a common platform and immunoassay, demonstrating high compatibility of biomarker trends across cohorts. After applying a pre-specified correction factor for α-synuclein, Aβ_40_, and Aβ_42_, (34), biomarker data were pooled across cohorts. CSF biomarker values outside of 3 times the interquartile range (IQR) below Q1 or above Q3 were defined as outliers and excluded from analysis (54/6930 values, 0.78%).

### MRI acquisition

In the ALFA+ cohort, a high-resolution 3D T1-weighted turbo field echo (TFE) sequence was acquired using a 3T Philips Ingenia CX scanner, with the following parameters: 0.75 mm isotropic voxel size, TR/TE of 9.9/4.6 ms, and a flip angle of 8°. For the WRAP and ADRC cohorts, all subjects were scanned on a GE 3T MR750 scanner equipped with a 32-channel head coil. The T1-weighted images in these cohorts were obtained using a 3D inversion recovery prepared fast spoiled gradient recalled (IR FSPGR) sequence with a 1 mm isotropic voxel size, TR/TE of 8.1/3.2 ms, and a flip angle of 12°, following a standardised protocol. Both protocols also included fluid-attenuated inversion recovery (FLAIR) scans to assess white matter hyperintensities.

The MRI scan performed the closest in time to the lumbar puncture was selected as the baseline scan (maximum difference of one year). A single follow-up MRI was conducted on average 3.5 years later (SD = 0.9), with a maximum interval of 6 years. The T1w image was segmented into GM, white matter (WM) and CSF tissue classes using SPM12. Total intracranial volume (TIV) was computed as the sum of GM, WM, and CSF volumes for each subject baseline image.

### MRI processing

Longitudinal analyses were performed using SPM12 (https://www.fil.ion.ucl.ac.uk/spm/software/spm12/) running in MATLAB (v2023b). All images were processed with pairwise longitudinal registration (PLR) to determine changes in GM volume over time. First, a non-linear registration between the baseline and coregistered follow-up T1w image was performed using high-dimensional deformations. This generated a whole-brain Jacobian determinant (JD) map for each participant, reflecting the amount of volumetric change per voxel between the two time-points. A negative difference score represents voxel contraction, whereas a positive difference score indicates volume expansion. Subject specific GM masks (> 0.2) were derived from the baseline T1w image in native space and applied to the JD maps. DARTEL normalisation was applied to standardise the JD maps to Montreal Neurological Institute (MNI) space. Finally, the normalised images were smoothed using an 8 mm Gaussian kernel. Additionally, given the central role of the hippocampus in AD, we quantified longitudinal change by computing the mean JD within a bilateral hippocampal mask using the AAL atlas.

### White matter hyperintensities

White matter hyperintensities (WMHs) were automatically segmented and quantified across all cohorts to derive total WMH lesion volume as the outcome measure. In the ALFA+ cohort, WMHs were automatically segmented using the BaMoS algorithm, an unsupervised algorithm based on a three-level Gaussian mixture model with Bayesian model selection, which incorporates both T1 and FLAIR images (35). For the ADRC and WRAP cohorts, WMHs were calculated using the Lesion Segmentation Tool (LST) in SPM12 (36). This lesion prediction algorithm uses both T1 and T2 FLAIR images to estimate the lesion probability at each voxel. These maps were then thresholded at a kappa of 0.5. The total WMH lesion volume from all participants was then normalized by dividing by the TIV, and data were log-transformed to resemble a normal distribution.

### Amyloid-PET

Amyloid PET scans were acquired on a Siemens Biograph mCT scanner, following a cranial computed tomography (CT) scan for attenuation correction, in the ALFA+ cohort. Four frames (4 × 5 min) were collected 90 to 110 min after the injection of 185 MBq [^18^F]flutemetamol (37). The images were reconstructed using an OSEM3D algorithm with eight iterations and 21 subsets, incorporating point spread function and time-of-flight corrections, into a 1.02 × 1.02 × 2.03 mm matrix. The averaged PET images were co-registered to the corresponding T1w MRI images and then normalised to MNI space using SPM12. The standardized uptake value ratio (SUVR) was calculated in MNI space using the standard target region (https://www.gaain.org/centiloid-project) with the whole cerebellum as a reference region. SUVR values were then transformed into the Centiloid (CL) scale using a previously calibrated conversion equation (38,39). For the WRAP and ADRC cohorts, [^11^C] Pittsburgh compound B (PiB) PET scans were performed using a Siemens EXACT HR+ scanner. Dynamic [^11^C]PiB PET scans were acquired for 70 minutes following a nominal 555-MBq injection. Distribution volume ratio (DVR) images were estimated using the cerebellar cortex as the reference region, and DRV values were linearly translocated to centiloids (40). Aβ-PET scans, performed at baseline, were available for a subset of participants (n = 405, 65%).

### Neuropsychological assessment

A modified version of the Preclinical Alzheimer Cognitive Composite (PACC) score was calculated, based on established protocols (41,42). For ALFA+, the PACC comprised (1) the Free and Cued Selective Reminding Test-Total immediate recall (43), (2) the WMS-IV Logical Memory delayed recall, (3) the WAIS-IV Coding, and (4) Animal Fluency (44). For WRAP/ADRC, the PACC consisted of (1) the Rey Auditory Verbal Learning Test, (2) the WMS-R Logical Memory delayed recall, (3) the WAIS-R Digit Symbol test, and (4) Animal Fluency (45).

All cognitive test scores were z-transformed using the baseline mean and standard deviation scores of A-T-N-participants (ALFA+: n=215; WRAP/ADRC: n=238) as a reference and then averaged for the PACC within cohort. The baseline test score for each individual was determined based on the date closest to the date of CSF collection. Neuropsychological test scores were collected over a follow-up period of up to 4.5 years (mean = 3.1, SD = 0.8), with participants completing an average of 2.6 cognitive assessments (SD=1.1).

### Statistical analyses

Comparisons of demographic characteristics between AT groups (*i.e.* A-T-, A+T-, A+T+) were performed using chi-squared tests for categorical variables and one-way ANOVA for continuous variables, with Tukey-adjusted pairwise comparisons of estimated marginal means when the omnibus test was significant.

#### Non-negative matrix factorization

The CSF NTK biomarkers, available at baseline, revealed highly correlated predictors based on a cross-correlation analysis among them (Supplementary Figure 1). Therefore, we implemented a Non-negative Matrix Factorization (NMF) approach to better understand the underlying structure of the CSF biomarker data. NMF is a data-driven method that can capture underlying non-linear patterns within complex datasets and is well equipped to deal with multicollinearity. NMF decomposes the matrix of CSF biomarkers into fundamental latent features, *i.e.* components of CSF biomarker combinations that represent unique patterns in the original data, via the product of two non-negative matrices *W* and *H* (Supplementary Figure 2). The rows in the *H* matrix provide the latent factors (*i.e.* biomarker components) by subject-specific coefficients, while the rows of the *W* matrix reflect the relative contribution of each biomarker to a latent factor (*i.e.* component). The component expression refers to the specific combination of predictor variables that make up the component and can be interpreted as “weights” indicating which CSF biomarkers contribute to each component and how they are combined. NMF applies non-negativity constraints, reflecting the non-negative nature of biomarker measures, enhancing the interpretability of results.

Prior to running NMF, multivariate imputation by chained equations (MICE) was applied to estimate missing CSF values (<1.2% of total) to avoid losing subjects (46). The imputation model used available CSF biomarker values of an individual along with age, sex, and *APOE ε4* carriership to accurately estimate a missing value; we used 50 imputations and 15 iterations per imputation. Subsequently, CSF biomarker values were normalised using min-max scaling to rescale values to a minimum of 0 and a maximum of 1. The Aβ_42/40_ ratio was inverted to align higher values with greater pathology for all biomarkers.

To determine the optimal number of components, NMF was run using different factorization ranks (*r*) from two to eleven (the total number of biomarkers). Given the stochastic nature of the NMF algorithm, the optimal factorization rank was selected as the best out of 50 runs with random initialisation. This procedure was then repeated using randomised data. For rank selection we considered the balance between high sparseness of the W matrix, quality of clustering (*i.e.*, low cophenetic dispersion), model fit (*i.e.* low residual sum of squares), and additional explained variance over a lower rank and randomised data (Supplementary Table 1), ensuring a balance between a good fit of the factorization and avoiding overfitting. After selection of the optimal number of components (*r*=6), the NMF was then refit at r = 6 with 200 runs. NMF analyses was performed using the *NMF* package in R (47).

For use in subsequent analyses, each participant had six component scores, with higher values indicating stronger expression of the biomarker pattern captured by that component. We first assessed the cross-sectional relationships between NMF-derived biomarker components with sex, age, *APOE ε4* carrier status, WMH, and AT stage via Pearson correlations, Welch t-tests, and one-way ANOVA with Tukey-adjusted contrasts.

#### Voxel-based morphometry

Next, second-level SPM multiple regression models related smoothed GM change maps to per-subject NMF biomarker component scores. For clarity, the MRI outcome was a whole-brain, voxel-wise JD rate map per-subject, reflecting within-subject local morphometric change (negative = contraction; positive = expansion) over the follow-up interval. We fitted: (i) single component models (one biomarker component as the predictor), and (ii) a joint all-components model (all six components entered simultaneously). Coefficients in (ii) represent unique partial effects conditional on the other components, since the effect of one component on GM volume changes may be influenced by the presence or absence of other components. Covariates in the models included age, sex, *APOE ε4* carrier status, baseline TIV, cohort, and interscan interval.

Both (i) and (ii) models were run in the full sample and repeated separately within A-T-, A+T-, and A+T+ groups to visualise possible stage-dependent spatial patterns. Design orthogonality and homogeneity were checked using CAT12. For all VBM analyses, the statistical significance threshold was set to *p* < 0.01 uncorrected to increase sensitivity to subtle changes expected in CU individuals over a short follow-up. Only clusters that survived an extent threshold of 100 voxels were considered.

#### Hippocampal ROI

Furthermore, to assess the association between longitudinal change in a hippocampal region of interest (ROI), an area highly vulnerable to AD pathology, and biomarker component expression, we performed linear regression: *mean hippocampus JD ∼ component + age + sex + APOE ε4 carrier status + TIV + cohort + interscan interval*. We used single-component models and the joint all-components model (all Variance inflation factor [VIF] <2). We additionally tested *component x AT* interactions and summarised AT-specific slopes with estimated trends.

#### Longitudinal cognition

Finally, the association of CSF components with cognitive trajectories was modelled with linear mixed models (LMM): *z-PACC ∼ time x component + age + sex + education + cohort + (11ID).* Where time is years since baseline, with random intercepts for subjects and fixed slopes were applied. In addition to examining the main effect of CSF component weight on cognitive decline across all participants, we stratified by amyloid status, since the large number of A-individuals may drive the overall association and mask a potential different relationship for individuals on the AD continuum. Furthermore, all models were repeated using the joint all-components model (all VIF<4). The LMM was performed using the lme4 package and visualised using ggeffects in R.

## Results

### Demographic characteristics

The study included 627 CU individuals with a mean age of 61.7 years (SD = 6.3). Of these, 62.0% were women (n = 389), and 46.7% were *APOE ε4* carriers (n = 289). The average education level was 14.9 years (SD = 3.3). Amyloid burden, as expressed in Centiloids, ranged from –19.9 to 155.4, with a mean value of 7.1 (Table 1).

**Table 1.** Subject characteristics.

|  | **A-T-** | **A+T-** | **A+T+** | **Total** |
| --- | --- | --- | --- | --- |
| n | 453 (72.2%) | 122 (19.5%) | 52 (8.3%) | 627 (100%) |
| Center, WADRC/WRAP/ALFA+ | 147/91/215 | 13/23/86 | 13/11/28 | 173/125/329 |
| Sex, f | 284 (62.7%) | 70 (57.4%) | 35 (67.3%) | 389 (62.0%) |
| Age, y | 60.7 (6.2)<sup>b,c</sup> | 63.2 (5.7)<sup>a,c</sup> | 66.4 (6.5)<sup>a,b</sup> | 61.7 (6.3) |
| Education, y | 15.2 (3.2)<sup>b,c</sup> | 14.3 (3.5)<sup>a</sup> | 14.0 (3.8)<sup>a</sup> | 14.9 (3.3) |
| Centiloid | -2.0 (7.3)<sup>b,c</sup> | 21.1 (28.3)<sup>a,c</sup> | 37.8 (35.0)<sup>a,b</sup> | 7.1 (22.7) |
| APOE ε4 carrier | 166 (36.7%)<sup>b,c</sup> | 94 (77.0%)<sup>a,c</sup> | 29 (55.8%)<sup>a,b</sup> | 289 (46.7%) |
| C1. Microglial reactivity | 0.14 (0.07)<sup>b</sup> | 0.09 (0.06)<sup>a,c</sup> | 0.13 (0.09)<sup>b</sup> | 0.13 (0.07) |
| C2. Cytokine signalling | 0.10 (0.06) | 0.10 (0.07) | 0.09 (0.07) | 0.10 (0.07) |
| C3. Neuroaxonal injury | 0.10 (0.07)<sup>b,c</sup> | 0.11 (0.06)<sup>a,c</sup> | 0.20 (0.09)<sup>a,b</sup> | 0.11 (0.08) |
| C4. Amyloid pathology | 0.09 (0.04)<sup>b,c</sup> | 0.22 (0.05)<sup>a,c</sup> | 0.27 (0.05)<sup>a,b</sup> | 0.13 (0.08) |
| C5. Tau-Synaptic injury | 0.11 (0.08)<sup>b,c</sup> | 0.14 (0.08)<sup>a,c</sup> | 0.38 (0.11)<sup>a,b</sup> | 0.14 (0.11) |
| C6. Astrocytic reactivity | 0.13 (0.08)<sup>c</sup> | 0.12 (0.08)<sup>c</sup> | 0.16 (0.10)<sup>a,b</sup> | 0.13 (0.08) |

Data are presented as mean (SD) or n (%); AT status was determined using a predetermined cohort specific cutoff for CSF Aβ42/40 ratio (A) and CSF p-tau181 (T); Centiloid values were available for n = 405 (65%) participants; f = female; y = years; APOE = Apolipoprotein E; a = significantly different from A-T-; b = significantly different from A+T-; c = significantly different from A+T+ at p<.05.

### Longitudinal changes in grey matter volume between AT stages

Longitudinal voxel-wise changes in GM volume showed distinct patterns between AT groups. Compared to A-T-, A+T-individuals demonstrated widespread atrophy, particularly in the medial temporal lobes, lateral temporal cortices, and frontal regions (Figure 1). Participants that were A+T+ showed subtle GM loss compared to A-T-. Clusters of GM increase were also observed in the occipital lobe. A+T+ individuals showed greater GM change than A+T-individuals in occipital and frontal cortices, which could reflect more increase or less decrease.

**Figure 1.**
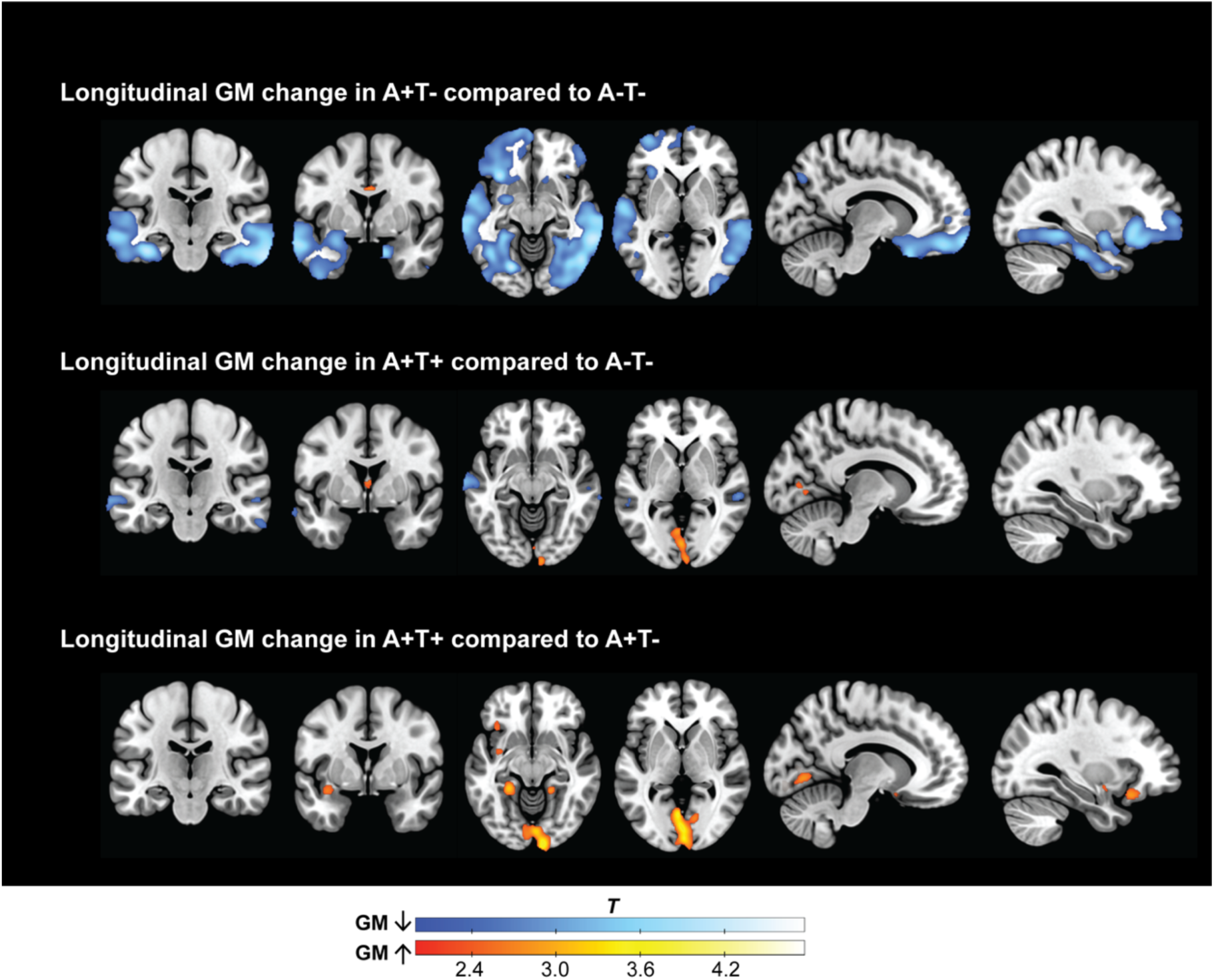
Longitudinal changes in grey matter volume across AT stages. Voxel-wise comparisons of GM volume changes over ± 3.5 years between AT stages. *Top)* A+T-compared to A-T-. *Middle)* A+T+ subjects compared to A-T-. *Bottom)* A+T+ subjects compared to A+T-. Blue indicates GM volume decreases, and red indicates GM volume increases in significant clusters relative to the comparison group. Threshold: *p* < 0.01, *k* > 100). All analyses are adjusted for age, sex, *APOE ϕ.4* carrier status, TIV, and time interval between scans.

### Non-negative matrix factorization (NMF)-derived biomarker components

NMF analysis identified six latent biomarker components that captured distinct patterns of CSF biomarker expression (Figure 2). The first component *(C1)* was characterised by strong loadings of sTREM2 (43%) and YKL-40 (25%), indicative of microglial and astrocytic reactivity, with a moderate co-expression of α-synuclein (18%). This component was significantly lower in A+T-individuals (Supplementary Figure 3), than in A-T-(mean difference= 0.055, 95% CI = [0.038, 0.072], *p*<.001) and lower than A+T+ individuals (0.043 [0.016, 0.070], *p*<.001). Lower levels of this component were also observed in *APOE ρ,4* carriers (t=5.62, *p*<.0001), and younger individuals (Spearman’s R=0.15, *p*<.001).

**Figure 2.**
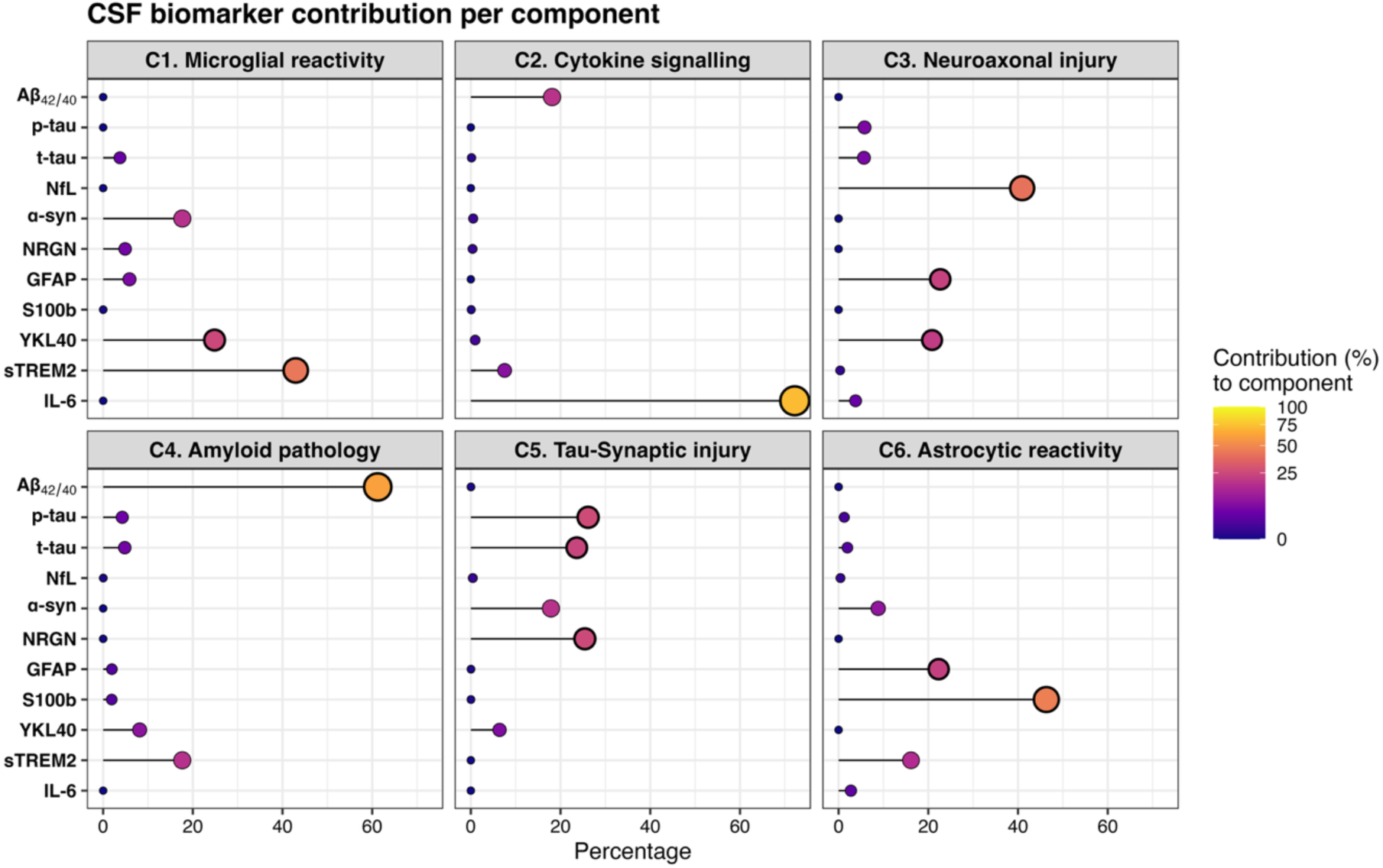
NMF-derived components. Lollipop plots showing the percentage contribution of each CSF biomarker to a non-negative matrix factorization (NMF) derived component. Aβ_42/40_ was inverted, higher scores reflect greater pathology. Components C1–C6 reflect latent patterns in CSF biomarker profiles across participants.

The second component *(C2)* was driven primarily by IL-6 (72%), with a moderate contribution from the Aβ_42/40_ ratio (18%). This cytokine-driven component showed no associations with amyloid or tau status, suggestive of non-AD related inflammatory processes. Higher levels of this component were associated with younger age (R=0.15, *p*<.001).

Component 3 *(C3)* was most strongly driven by NfL (41%), a marker of neuroaxonal injury, and showed moderate co-expression of astrocytic biomarkers GFAP (23%) and YKL-40 (21%). This component expression increased strongly with age (R=0.60, *p*<.001), and larger WMH (R=0.13, *p*<.01), and was more frequently observed in males (t=4.55, *p*<.0001) and demonstrated a stepwise increase across AT stages. A+T– individuals had higher expression levels than A–T–(mean difference = 0.017, 95 % CI [0.000, 0.033], *p* < 0.05), A+T+ individuals showed higher levels than both A–T– (0.105 [0.080, 0.129], *p* < 0.001), and A+T– (0.087 [0.059, 0.115], *p* < 0.001). The fourth component *(C4)* reflected Aβ pathology (61%), with high loadings of the Aβ_42/40_ ratio and moderate sTREM2 expression (18%). As expected, this component was expressed strongly in A+ individuals, A+T-had higher C4 levels than A-T-(mean difference = 0.135, 95% CI = [0.125, 0.146], *p*<.0001), and A+T+ was higher than A-T-(0.182 [0.166, 0.197], *p*<.0001), with even higher levels in A+T+ relative to A+T-(0.046 [0.029, 0.064], *p*<.0001). This component also correlated positively with older age (R=0.29, *p*<.001), larger WMH (R=0.19, *p*<.001), and had a higher expression in *APOE ρ,4* carriers (t=-8.45, *p*<.0001).

Component 5 *(C5)* captured a profile of tau-related pathophysiology and synaptic injury. It showed high loadings of p-tau (26%), t-tau (24%), α-synuclein (18%), and neurogranin (25%). Expression of this component increased with age (R=0.22, *p*<.001) and was slightly higher in females (t=-2.21, *p*<.05). Moreover, it was significantly elevated in both A+T-(mean difference 0.025, 95% CI = [0.004, 0.045], *p*<.05), and A+T+ (0.270 [0.241, 0.300], *p*<.0001) compared to A-T-, with the highest expression in the A+T+ group compared to A+T-(0.245 [0.212, 0.278], *p*<.0001).

Finally, component 6 *(C6)* represented astrocytic reactivity, mainly represented by s100B (46%) and GFAP (22%). This component was elevated in A+T+ individuals relative to both A+T-(mean difference = 0.039, 95% CI = [0.008, 0.071], *p*<.05), and to A-T-individuals (0.029 [0.001, 0.057], *p*<.05). Expression of this component was negatively correlated with WMH load (R=-0.16, *p*<.001) and was slightly higher in males (t=3.05, *p*<.01).

### Biomarker component and longitudinal grey matter changes

First, we investigated the association of single biomarker components and GM volume change. Higher expression of the *Microglial Reactivity* component (C1) was associated with subtle regional GM changes in both directions, *i.e.* volume decreases and increases (Figure 3). These associations became more pronounced when stratified by AT stage. In A+T-individuals, higher C1 was linked to volume increases in the temporal pole, parahippocampal gyrus, and fusiform gyrus, while in individuals with tau pathology, *i.e.* A+T+, elevated C1 was associated with neuronal loss in the orbitofrontal cortex, hippocampus, inferior temporal gyrus, and cingulate cortex. This AT stage dependent relationship was also reflected by the ROI-based analysis, which showed a positive association of C1 with hippocampal volume change in A+T-, and a negative association in A+T+ individuals, although only in the larger sample of A-T-this relationship was significant (Figure 3).

**Figure 3.**
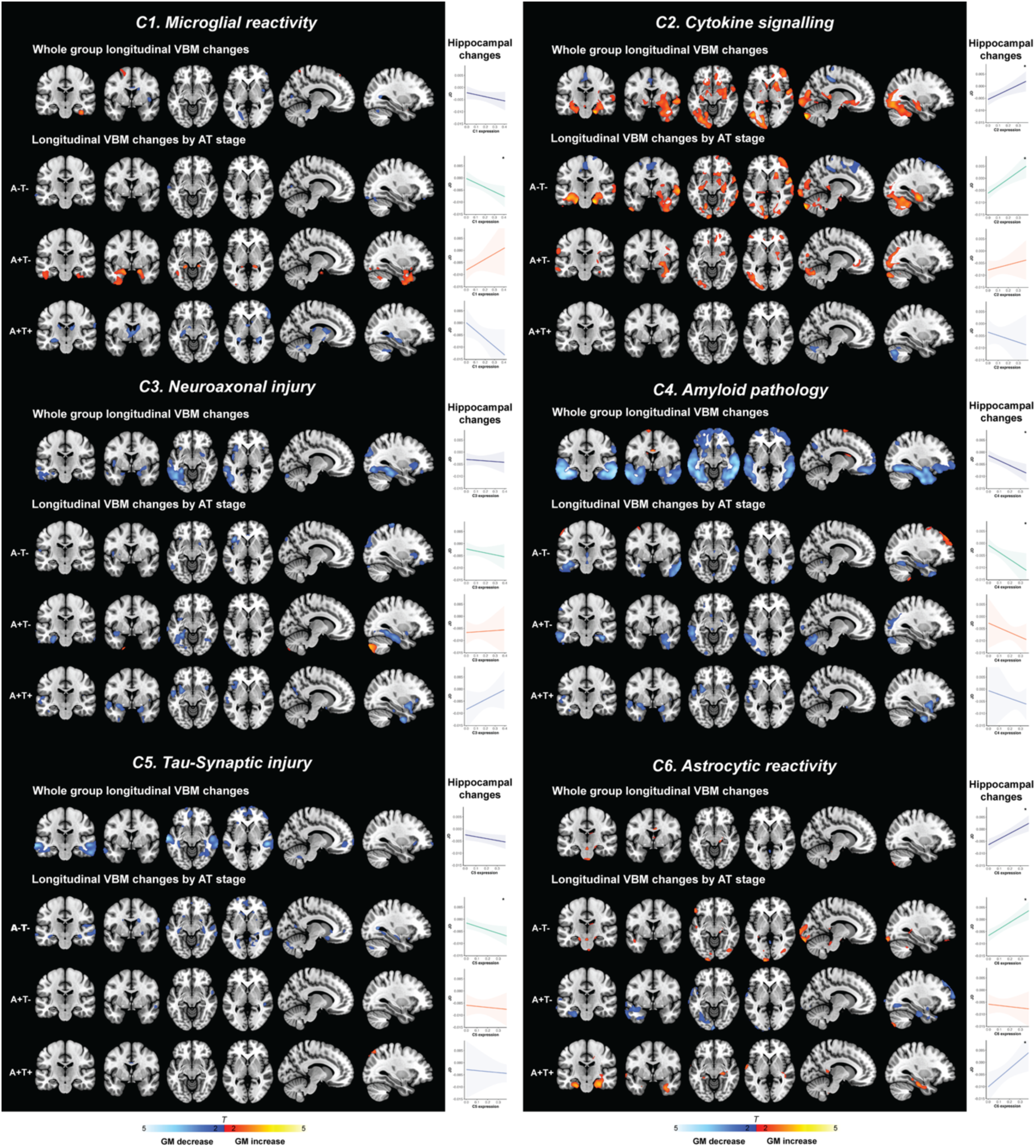
Voxel-wise associations of longitudinal grey matter volume changes with CSF biomarker components. Overview of different biomarker components and its association with longitudinal grey-matter changes. Voxel-wise associations of longitudinal grey matter volume changes (*i.e.* Jacobian determinants [JD]) with biomarker components (C1-C6) in the entire sample and by AT stage. Brain maps display regions where higher expression of components are associated with significant GM volume decreases (blue) or increases (red). Threshold: *p* < 0.01, *k* > 100. Complementing plots show the association between component (C1-C6) expression and longitudinal hippocampal change in the entire sample, and within AT subgroups. All analyses are adjusted for age, sex, *APOE ϕ.4* carrier status, TIV, and time interval between scans. Significance level: * *p* < .05.

Higher levels of C2, predominantly reflecting IL-6 related inflammation, were associated with widespread GM volume increases in temporal and occipital regions, including the hippocampus, amygdala, parahippocampal gyrus, superior and middle temporal gyri, temporal pole, cerebellum, middle occipital gyrus, and medial frontal cortex (Figure 3). This cytokine-related hypertrophy was not observed in A+T+ individuals.

Elevated levels of neuroaxonal injury and reactive astrocytes (C3) were associated with GM atrophy in the inferior and middle temporal gyri, fusiform gyrus, and inferior occipital cortex across all AT stages (Figure 2). The *Aβ Pathology* component (C4) was robustly associated with widespread atrophy, including the inferior and middle temporal gyri, fusiform area, temporal pole, inferior occipital gyrus, and orbitofrontal cortex. This was observed even in the A-T-group. Expression of *Tau-Synaptic Injury* (C5) was related to neuronal loss in the fusiform gyrus, parahippocampal gyrus, inferior temporal gyrus, inferior occipital cortex, precuneus, and orbitofrontal cortex.

The *Astrocytic Reactivity* component (C6) showed both subtle hypertrophic and atrophic associations with GM volume, which varied by AT stage. In A-T-individuals, higher C6 expression was associated with volume increases in the nucleus accumbens, anterior cingulate cortex, and inferior occipital cortex. In contrast, in A+T-individuals, higher C6 levels were associated with atrophy in the fusiform gyrus, inferior occipital cortex, and orbitofrontal cortex. In A+T+ individuals, astrocyte-related hypertrophy was observed in the parahippocampal gyri, fusiform gyrus, and posterior cingulate cortex. These diverging stage-dependent effects were demonstrated as well by the positive associations of C6 levels and hippocampal change in A-T-and A+T+ individuals, but not in A+T-(Figure 3).

### Joint contributions of biomarker component to longitudinal grey matter changes

To examine the unique association of each biomarker component with GM volume change, we fitted a joint model that included all components simultaneously (Supplementary Figure 4). Importantly, in this model, *Aβ Pathology* (C4) remained strongly associated with widespread atrophy, indicating a robust association after accounting for the other biological processes. Similarly, *Microglial Reactivity* (C1), *Cytokine Signalling* (C2), and *Astrocytic Reactivity* (C6) retained similar significant associations with GM change, suggesting associations that are not explained by the other components in the model. In contrast, the *Neuroaxonal Injury* component (C3) showed attenuated effects, and *Tau-Synaptic Injury* (C5) was no longer significantly associated with GM volume change. This suggests that the initial observed C5 effect on neuronal loss may be partly attributed to shared variance with Aβ pathology and axonal injury, rather than a distinct effect.

### Biomarker components and cognition

Higher expression of the *Aβ Pathology* component (C4) was associated with reduced performance over time on the PACC composite (β±SE = –0.45±0.07, *p*<.0001; Supplementary Figure 5). Furthermore, higher levels of *Microglial Reactivity* (C1) and *Astrocytic Reactivity* (C6) were associated with better cognitive performance over time (C1: β±SE = 0.34±0.08, *p*<.0001; C6: β±SE = 0.19±0.07, *p*<.01). When stratified by Aβ status, the positive association between microglial and astroglial component levels and cognition was present in Aβ-negative controls (C1: β±SE = 0.26±0.10, *p*<.01; C6: β±SE = 0.26±0.08, *p*<.01) but was no longer observed in individuals on the AD continuum. All these relationships remained significant after adjusting for the other biomarker components in the same model (Supplementary Figure 6). No significant associations with cognitive change were observed for the *Cytokine Signalling* (C2), *Axonal Injury* (C3), or *Tau-Synaptic Injury* (C5) components, regardless of Aβ status.

## Discussion

In this large longitudinal multi-cohort study of CU individuals across different stages of the AT continuum, we investigated the biological processes underlying GM volume changes over time. To address this, we identified data-driven CSF biomarker components of different pathophysiological pathways involved in the biological progression of AD and investigated their associations with longitudinal changes in GM volume and cognitive performance. Using NMF, we derived distinct biomarker profiles indicative of Aβ pathology, tau-related pathophysiology with synaptic injury, neuroaxonal injury, astroglial and microglial reactivity, and cytokine signalling. Our main findings were that Aβ pathology was the strongest correlate of early GM atrophy, independent of tau pathophysiology or neuroaxonal injury. In addition, we found that glial and inflammatory responses are associated with longitudinal increases in GM volume, with effects varying by AT stages. A better understanding of these complex dynamic relationships between structural brain changes and diverse biological pathways at the earliest stages of AD is crucial to inform the development of interventions before irreversible neurodegeneration occurs.

The strongest and most widespread GM atrophy was associated with the *Aβ Pathology* component (C4), after mutual adjustment of tau-related pathophysiology, neuroaxonal injury, and other biomarker processes. In line with this, A+T-individuals already showed significant GM loss compared to A-T-individuals, predominantly in temporal and frontal regions, and higher Aβ pathology was associated with worse cognitive performance over time. These observations are consistent with recent work (12,48) emphasizing that early Aβ accumulation is neither a benign process nor solely a triggering event in the AD cascade. Rather, recent studies suggest that early Aβ pathology has direct neurotoxic effects, causing neuronal changes and cognitive impairment prior to significant tau deposition and clinical symptoms (23,49–54). Furthermore, our results align with previous reports suggesting that Aβ and tau pathology, in addition to their synergistic effects, may also contribute independently to neuronal dysfunction, especially in early disease stages (55).

We found that higher expression of the *Tau-Synaptic Injury* component (C5) was initially associated with GM volume loss in medial and lateral temporal as well as frontal regions. However, this association did not remain significant after adjusting for the effects of other biomarker components, suggesting a shared variance at these early stages of the disease. These findings contrast with the dominant view that tau pathology is the primary driver of neurodegeneration, and that Aβ correlates rather weakly with atrophy (5,56,57), and that the presence of tau tangles is necessary in A+ individuals to observe atrophy (3,20,58,59). Notably, this view is largely based on studies conducted in symptomatic stages of AD, where tau pathology is more advanced and more strongly correlated with GM atrophy, while Aβ levels typically reached a plateau and show little correlation with atrophy (60,61). In contrast, our results suggest that these relationships may differ in the very earliest stages of the disease from later stages. In preclinical stages, the dynamic range of tau pathology is lower, and CSF p-tau may primarily reflect soluble phosphorylated tau species rather than fibrillar aggregates (62). These soluble tau forms may have less pronounced effects than Aβ on GM atrophy, at this preclinical stage. Moreover, it should be noted that the biomarker components were derived using a data-driven approach. Therefore, the way variance is partitioned across components may reflect more statistical than purely biologically separability.

Our observations support the rationale for anti-amyloid therapies early in the disease course, before extensive neuronal loss and significant tau pathology occurs. Illustrative, current ongoing trials of lecanemab and donanemab are targeting asymptomatic individuals (63,64). Intervening at this preclinical stage may be more likely to lead to better clinical outcomes than when initiated later in the disease.

In contrast to the associations of core AD pathology biomarkers with atrophy, we observed that components reflecting increased microglial and astrocytic reactivity, as well as elevated cytokine response, were independently associated with longitudinal increases in GM volume in a stage-dependent manner. Higher expression of the *Microglial Reactivity* component (C1), characterised by strong loadings of sTREM2 and YKL-40, showed independent associations with GM volume increases in A+T-individuals especially in temporal areas, whereas in A+T+ individuals higher C1 levels correlated with GM volume loss. Similarly, the *Astrocytic Reactivity* component (C6), characterised by high expression of S100B and GFAP, showed an AT-stage dependent relationship with GM changes. Associating with hypertrophy in A-T– and A+T+ individuals but linked to atrophy in A+T-individuals. Moreover, the *Cytokine Signalling* component (C2), predominantly driven by IL-6, showed widespread associations with GM volume increases across temporal and occipital regions. No differences in expression levels of this component were observed between AT groups, suggesting a contribution of non-AD related inflammatory processes.

Several prior studies have reported paradoxical increases in GM volume in individuals with preclinical AD, rather than the expected atrophy characteristic of a neurodegenerative disease (13–18,20,65–69). While initially counterintuitive, this observation has been consistently reported across multiple independent cohorts, suggesting that it is not merely a methodological artifact. However, the underlying biological substrates driving these volume increases remained uncertain.

Our results provide the first large longitudinal *in vivo* evidence indicating neuroinflammatory processes as a potential contributor to this observation in CU individuals. This may include cellular processes such as gliosis, inflammation induced swelling, and compensatory processes in response to AD pathology (66,70–74). Specifically, the enlargement of cell bodies, which may be from microglia and astrocyte activation and recruitment, as well as neuronal hypertrophy in response to early AD pathology, could underlie this observation (75,76). Supporting this, increased glucose metabolism in preclinical AD has been described in previous research (18,40), potentially reflecting increased glial activity before overt neurodegeneration.

Our observation that volume increases were stage-dependent further supports the notion that glial reactivity may play both protective and harmful roles and how a non-linear expression depending on the disease phase (77–81). Additionally, the use of longitudinal MRI and cognitive trajectories in this study was critical for detecting these changes, as cross-sectional studies may overlook dynamic or non-linear structural trajectories (8,16,19,20), and have interpreted thicker cortices as indicative of reserve or resilience (17,82,83). We observed that higher expression of microglial and astrocytic reactivity components was associated with better cognitive outcomes over time. When stratifying by Aβ status this association was observed only in individuals without Aβ pathology, suggesting that the whole sample analysis was driven by the A-subgroup, and that initially a homeostatic or protective glial response may support cognitive resilience, but that once AD-pathology accumulates this response becomes less effective or contributes to downstream neurodegeneration.

Furthermore, our findings have potential implications for interpreting recent findings from amyloid immunotherapy trials, where paradoxical GM volume loss was reported when amyloid burden was lowered, which was associated with beneficial cognitive outcomes (84,85). This observation has generated considerable debate and raised concerns about possible neurotoxic effects (86,87). However, our results support the hypothesis that this loss of brain volume may arise from beneficial anti-inflammatory effects. Specifically, Aβ pathology might provoke neuroinflammation-associated swelling of brain tissue, which then following amyloid removal, manifests as volume reduction. Indicating that this pseudoatrophy does not reflect greater neurodegeneration, nor relates to worse cognitive outcomes (88).

Strengths of this study include its large sample size, the use of longitudinal MRI, which remains limited in preclinical AD research, and the harmonised CSF biomarker data across cohorts using the same assay. This allowed us to more accurately detect dynamic and potentially non-linear structural changes. Furthermore, our NMF approach enabled us to capture biologically meaningful patterns in CSF biomarkers that naturally co-occur, and to disentangle their overlapping contributions to GM changes. In contrast, prior studies often analysed individual biomarker associations in isolation, ignoring inter-correlations or artificially splitting shared variance into separate effects, which may be mathematically arbitrary and biologically less meaningful. Moreover, this study incorporates biomarkers beyond core AD pathology, offering better insight into the complex multifactorial nature of early AD. Limitations include potential variability introduced from the use of two different MRI scanners and protocols, although to mitigate this effect cohort was included as a covariate in all VBM analysis. Moreover, given the very early disease stage of our participants, relatively low tau pathology was present, which may limit generalisability into more advanced and symptomatic AD stages. Furthermore, it must be recognised that our study participants come from specific research cohorts with strict inclusion criteria, and our findings should be replicated in a more diverse population.

## Conclusions

This study provides new insight into the biological mechanisms underlying longitudinal GM changes at the earliest stages of AD. We demonstrate that at the earliest asymptomatic stages of the AD continuum, Aβ pathology is strongly associated with GM atrophy and predictive of cognitive decline, independent of tau pathophysiology and neuroaxonal injury, supporting preventive anti-amyloid interventions. Furthermore, glial reactivity and cytokine related processes are associated with GM volume increases in a stage-dependent manner, suggesting a role of neuroinflammation in shaping structural brain changes.

As volumetric MRI is used as a secondary outcome measure in clinical trials, and those trials are moving to earlier disease stages, it is essential to understand which factors contribute to these non-linear, and often paradoxical, changes. Our findings illustrate the complexity and heterogeneity of AD. Future investigation should therefore aim for even more fine-grained longitudinal studies, for example by incorporating novel astro– and microglial PET and proteomic analyses, to ultimately achieve a more precise understanding of early AD pathophysiology. Ultimately, a better understanding of these early biological processes will improve identification of therapeutic targets and inform the design of more personalised preventative strategies at the earliest, and likely most modifiable, stages of the disease.

## Declarations

### Ethics approval and consent to participate

The ALFA+ study (ALFA-FPM-0311) was approved by the Independent Ethics Committee ‘Parc de Salut Mar’, Barcelona, and registered at Clinicaltrials.gov (Identifier: NCT02485730). All participating subjects signed the study’s informed consent form, which was approved by the Independent Ethics Committee ‘Parc de Salut Mar’, Barcelona. The Wisconsin Alzheimer’s Disease Research Center – Clinical Core Study and the Wisconsin Registry for Alzheimer’s Prevention were approved by the University of Wisconsin Madison Health Sciences Institutional Review Board. The studies were conducted according to the Declaration of Helsinki.

### Consent for publication

Not applicable

### Availability of data and materials

All requests for raw and analysed data and materials will be promptly reviewed by the corresponding authors and the Barcelonaβeta Brain Research Center to verify whether the request is subject to any intellectual property or confidentiality obligations. Bulk Anonymized data can be shared by request from any qualified investigator for the sole purpose of replicating procedures and results presented in the article, providing data transfer is in agreement with EU legislation and decisions by the Institutional Review Board (IRB) of each participating center. For any other purposes, the data request will be processed by internal data request procedures of the participating institute(s).

Data utilized in the WRAP cohort consist of sensitive, human research participant data and participants have not signed informed consent to have their data shared in public repositories for publications. Therefore, data deposition is unethical. Data are available upon request for authorized researchers who meet the criteria for access to confidential data.

### Competing interests

W.P., R.C, M.K., O.G-R., C.M, R.E.W., E.M.J., and R.E.L. declare no competing interests. J.L.M. is currently a full-time employee of H. Lundbeck A/S and previously has served as a consultant or on advisory boards for the following for-profit companies or has given lectures in symposia sponsored by the following for-profit companies: Roche Diagnostics, Genentech, Novartis, Lundbeck, Oryzon, Biogen, Lilly, Janssen, Green Valley, MSD, Eisai, Alector, BioCross, GE Healthcare, and ProMIS Neurosciences. G.K. is a full-time employee of Roche Diagnostics GmbH, Penzberg, Germany. C.Q-R. is a full-time employee of Roche Diagnostics International Ltd, Rotkreuz, Switzerland. K.B. has served as a consultant and at advisory boards for Abbvie, AC Immune, ALZPath, AriBio, Beckman-Coulter, BioArctic, Biogen, Eisai, Lilly, Moleac Pte. Ltd, Neurimmune, Novartis, Ono Pharma, Prothena, Quanterix, Roche Diagnostics, Sunbird Bio, Sanofi and Siemens Healthineers; has served at data monitoring committees for Julius Clinical and Novartis; has given lectures, produced educational materials and participated in educational programs for AC Immune, Biogen, Celdara Medical, Eisai and Roche Diagnostics; and is a co-founder of Brain Biomarker Solutions in Gothenburg AB (BBS), which is a part of the GU Ventures Incubator Program, outside the work presented in this paper. H.Z. has served on scientific advisory boards and/or as a consultant for Abbvie, Acumen, Alector, Alzinova, ALZPath, Annexon, Apellis, Artery Therapeutics, AZTherapies, Cognito Therapeutics, CogRx, Denali, Eisai, Nervgen, Novo Nordisk, Optoceutics, Passage Bio, Pinteon Therapeutics, Prothena, Red Abbey Labs, reMYND, Roche, Samumed, Siemens Healthineers, Triplet Therapeutics, and Wave; has given lectures in symposia sponsored by Cellectricon, Fujirebio, Alzecure, Biogen, and Roche; and is a co-founder of Brain Biomarker Solutions in Gothenburg AB (BBS), which is a part of the GU Ventures Incubator Program (outside submitted work). M.S-C. has received in the past 36mo consultancy/speaker fees (paid to the institution) from by Almirall, Biogen, Beckman Coulter, Eli Lilly, Quanterix, Novo Nordisk, and Roche Diagnostics. He has received consultancy fees or served on advisory boards (paid to the institution) of Eli Lilly, Grifols, Novo Nordisk, and Roche Diagnostics. He was granted a project and is a site investigator of a clinical trial (funded to the institution) by Roche Diagnostics. In-kind support for research (to the institution) was received from ADx Neurosciences, Alamar Biosciences, ALZpath, Avid Radiopharmaceuticals, Eli Lilly, Fujirebio, Janssen Research & Development, Meso Scale Discovery, and Roche Diagnostics; M.S-C. did not receive any personal compensation from these organizations or any other for-profit organization. G.S. has received speaker fees from Springer, GE Healthcare, Biogen, Esteve and Adium and advisory fees from Johnson&Johnson. S.C.J. has served as an advisor to Ely Lilly, Enigma Biomedical, AlzPath, Alamar and Sunbird Bio. J.D.G. is currently a full-time employee at AstraZeneca, and has received research support from GE Healthcare, Roche Diagnostics, Hoffmann – La Roche, Life-Molecular Imaging and Philips Netherlands, speaker/consulting fees from Biogen, Roche Diagnostics, Life-Molecular Imaging and Esteve, participated in the Molecular Neuroimaging Scientific Advisory Board of Prothena Biosciences, and is the founder and co-owner of BetaScreen SL.

### Funding

The ALFA+ study receives funding from “la Caixa” Foundation, under agreement LCF/PR/SC22/68000001 and the Alzheimer’s Association and an international anonymous charity foundation through the TriBEKa Imaging Platform project (TriBEKa17519007). Additional support has been received from the Universities and Research Secretariat, Ministry of Business and Knowledge of the Catalan Government under the grant no. 2021 SGR 00913. The Wisconsin Registry for Alzheimer’s Prevention is funded by NIH AG027161 and the Wisconsin ADRC by AG062715. Additional NIH funding for CSF collection and analysis was provided by AG021155, AG037639, and AG062167. W.P. is supported by an Early Career Grant – Call Biomedical Research by Alzheimer Nederland (WE.03-2025-05). R.C. receives support from the grant RYC2021-031128-I, funded by MCIN/AEI/10.13039/501100011033 and the European Union NextGenerationEU/PRTR. O.G-R. receives funding from the Alzheimer’s Association Research Fellowship Program (2019-AARF-644568), from Instituto de Salud Carlos III (ISCIII) through the project PI19/00117 co-funded by the European Union (FEDER), and from Spanish Ministry of Science and Innovation – State Research Agency MCIN/AEI/10.13039/501100011033 through the project IJC2020-043417-I, co-funded by the European Union “Next GenerationEU”/PRTR. M.S-C. receives funding from the European Research Council (ERC) under the European Union’s Horizon 2020 research and innovation programme (Grant agreement No. 948677); ERA PerMed-ERA NET and the Generalitat de Catalunya (Departament de Salut) through the project SLD077/21/000001; Project “PI22/00456, funded by Instituto de Salud Carlos III (ISCIII) and co-funded by the European Union; and from a fellowship from “la Caixa” Foundation (ID 100010434) and from the European Union’s Horizon 2020 research and innovation programme under the Marie Skłodowska-Curie grant agreement No 847648 (LCF/BQ/PR21/11840004). G.S. received funding from the European Union’s Horizon 2020 Research and Innovation Program under Marie Sklodowska-Curie action grant agreement number 101061836, an Alzheimer’s Association Research Fellowship (AARF-22-972612), the Brightfocus Foundation (A2024007F), the Alzheimerfonden (AF-980942, AF-994514, AF-1012218), Greta och Johan Kocks research grants and travel grants from the Strategic Research Area MultiPark (Multidisciplinary Research in Parkinson’s Disease) at Lund University, and from the ADDF at BBRC.

### Authors’ contributions

W.P. and J.D.G. conceptualised the study. W.P. performed the data analysis, created the figures, and wrote the manuscript. G.S. and J.D.G supervised the project and provided revisions. R.C. and M.K. contributed expertise on neuroimaging analysis. R.C., M.K., O.G.R, C.M., J.L.M., G.K, C.Q-R., R.E.W., M.S.C., and S.C.J. contributed to the interpretation of findings and provided feedback on the manuscript.

## Data Availability

All requests for raw and analysed data and materials will be promptly reviewed by the corresponding authors and the BarcelonaBeta Brain Research Center to verify whether the request is subject to any intellectual property or confidentiality obligations. Bulk Anonymized data can be shared by request from any qualified investigator for the sole purpose of replicating procedures and results presented in the article, providing data transfer is in agreement with EU legislation and decisions by the Institutional Review Board (IRB) of each participating center. For any other purposes, the data request will be processed by internal data request procedures of the participating institute(s).
Data utilized in the WRAP cohort consist of sensitive, human research participant data and participants have not signed informed consent to have their data shared in public repositories for publications. Therefore, data deposition is unethical. Data are available upon request for authorized researchers who meet the criteria for access to confidential data.

## List of abbreviations

A: Amyloid
AAL: Automated Anatomical Labeling
Aβ: amyloid-beta
AD: Alzheimer’s disease
ADRC: Wisconsin Alzheimer’s Disease Research Center
ALFA: For ALzheimer’s and Families
APOE: apolipoprotein E
α-syn: α-synuclein
C: Component
CDR: Clinical Dementia Rating
CL: Centiloid
CSF: cerebrospinal fluid
CT: computed tomography
CU: cognitively unimpaired
DVR: distribution volume ratio
GFAP: glial fibrillary acidic protein
GM: grey matter
IL-6: interleukin-6
IQR: interquartile range
JD: Jacobian determinant
LMM: linear mixed models
MICE: multivariate imputation by chained equations
MNI: Montreal Neurological Institute
MRI: magnetic resonance imaging
NfL: neurofilament light protein
NMF: non-negative matrix factorization
NRGN: neurogranin
NTK: NeuroToolKit
PACC: Preclinical Alzheimer Cognitive Composite
p-tau: phospho-tau
PLR: pairwise longitudinal registration
S100B: S100 calcium binding protein B
sTREM2: soluble triggering receptor expressed on myeloid cells 2
SUVR: standardized uptake value ratio
T: Tau
t-tau: total-tau
TIV: Total intracranial volume
VBM: voxel-based morphometry
VIF: variance inflation factor
WM: white matter
WMHs: white matter hyperintensities
WRAP: Wisconsin Registry for Alzheimer’s Prevention
YKL-40: chitinase-3-like protein 1

## Acknowledgements

This publication is part of the ALFA study (ALzheimer and FAmilies). The authors would like to express their most sincere gratitude to the ALFA project participants and relatives without whom this research would not have been possible. Collaborators of the ALFA study are: Aida Fernandez, Aitana Plaza, Alba Cañas, Alba Martos Sánchez, Alba Fernández Bonet, Albert Rodrigo-Pares, Albina Polo, Aldana Lizarraga, Aleix Puig, Ana Fernández-Arcos, Ana Harris, Andrea Ambite, Andreea Rădoi, Anna Brugulat-Serrat, Anna Coward, Anna Soteras, Annabella Beteta, Arcadi Navarro, Armand González-Escalante, Bet Travesset Muntada, Blanca Rodríguez-Fernández, Carles Falcón, Carlota Medina, Carme Deulofeu, Clara Abadías, Clara Gallay, Claudia Porta-Mas, Claudia Vasallo, Cristina Mustata, David Fusté, David López-Martos, David Vállez, Diana Palacios, Diego Cascales, Eider Arenaza Urquijo, Eleni Palpatzis, Elisabet Zhan Travesset Muntada, Elisabeth Ferrer i Mairal, Esther Jiménez, Eva Palacios, Federica Anastasi, Felipe Hernández-Villamizar, Fernanda Campos Strazzi, Fernando Gaston Rossi, Ferran Lugo, Francisco Javier Melendez, Gema Huesa, Gianmarco Iaccarino, Gonzalo Sánchez-Benavides, Grégory Operto, Helena Blasco, Iman Sadeghi, Irene Cumplido, Irene Navalpotro, Isabel Estragués, Isabel Perez, Israel Turull, Iva Knezevic, Jaume Roca Alcaraz, Javier Torres-Torronteras, Jordi Camí, Jordi Freixa, Jordi Huguet, Jordi Peña-Casanova, José Contador, José María González de Echávarri, Kahina Baouche, Karine Fauria, Laia Tenas, Laura Gusó, Laura Hernandez, Laura Iglesias, Laura Stankeviciute, Leydi Dayana Martinez, Lidia Canals Gispert, Lluis Solsona, Mahnaz Shekari, Maite Gandia Ferrero, Manuel Garfia, Marc Vilanova, Marco Bianchi, Maria Emilio, Maria León, Maria Roman, Maria Pujol-Torrens, Marina de Diego, Marina García, Marta Crous Bou, Marta del Campo, Marta Félez, Marta Milà Alomà, Mireia Sánchez, Montserrat Vilà, Müge Akinci, Natalia Vilor-Tejedor, Neus de la Cruz-Sanchez, Nina Gramunt Fombuena, Noe Rodriguez de Guzmán Gallego, Noelia Giselle Rugna, Noelia Rodríguez de Guzmán Gallego, Núria Lleonart I Camps, Núria Tort Colet, Patricia Genius, Pau Sánchez, Paula Marne, Paula Ortiz, Pauline Diana Martens, Pilar Tartière-González, Rafael Dal-Ré, Ricardo Aquite, Ricardo Berbería, Ruth Dominguez, Ruth Pareja Alcaraz, Sabrina Segundo, Sandra Pradas, Sara Aragó, Sarata Sall Sall, Sherezade Fuentes, Tania Menchón, Tavia Evans, Vasiliki Bikou, Xavi Meléndez, Xavier Gotsens. The authors thank Roche Diagnostics International Ltd for providing the kits to measure CSF biomarkers, and the laboratory technicians at the Clinical Neurochemistry Lab in Mölndal, Sweden, who performed the analyses. The NeuroToolKit is a panel of exploratory prototype assays designed to robustly evaluate biomarkers associated with key pathologic events characteristic of AD and other neurological disorders, used for research purposes only and not approved for clinical use (Roche Diagnostics International Ltd, Rotkreuz, Switzerland). We would like to thank GE healthcare for kindly providing [^18^F]flutemetamol doses of ALFA+ participants.

## Notes

### Clinical Trial

NCT01835717

### Author Declarations

The Independent Ethics Committee ‘Parc de Salut Mar’, Barcelona, gave ethical approval for the ALFA+ study (ALFA-FPM-0311). The University of Wisconsin Madison Health Sciences Institutional Review Board gave ethical approval for the Wisconsin Alzheimer's Disease Research Center - Clinical Core Study and the Wisconsin Registry for Alzheimer's Prevention.

