## Supplementary material for "Neurobiological correlates of longitudinal grey matter volume changes in preclinical Alzheimer’s disease"

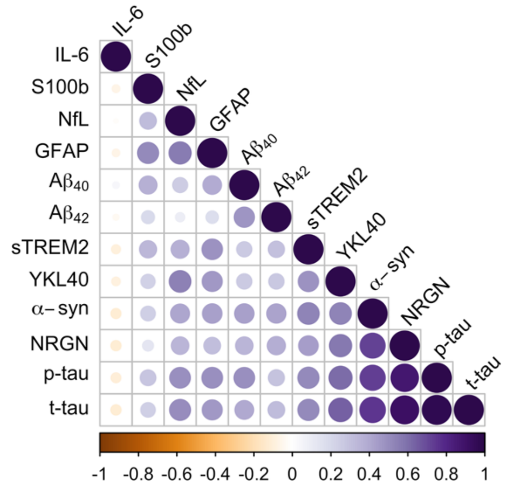


**Supplementary Figure 1. Correlation matrix of CSF biomarkers**

Spearmans's rank correlation coefficients between individual CSF biomarkers. Variables are hierarchically clustered. Colour scale shows correlation strength and direction. Only significant correlations ($p < .05$) are shown. Abbreviations: Aβ = amyloid-β; GFAP = glial fibrillary acidic protein; IL-6 = interleukin-6; NfL = neurofilament light; p-tau = phosphorylated tau; S100b = S100 calcium binding protein B; sTREM2 = soluble triggering receptor on myeloid cells 2; t-tau = total tau.

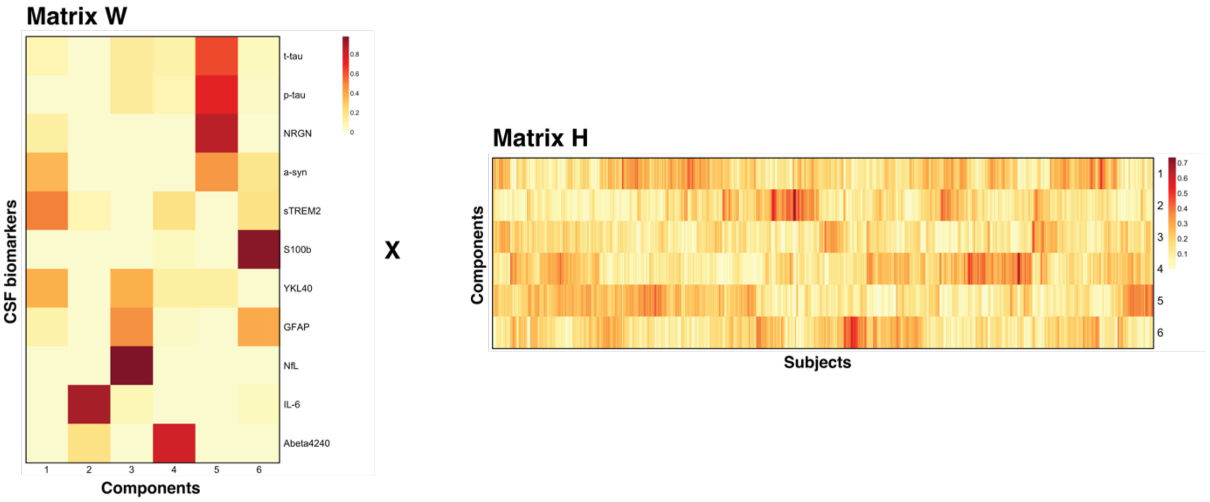


**Supplementary Figure 2. Non-negative matrix factorization approach**

The original data, represented as a matrix of CSF biomarkers values (*rows*) by subject (*column*), is approximated by the product of matrices W and H. Matrix W captures the contribution of each biomarker (*rows*) to components (*columns*), while Matrix H reflects the component expression (*rows*) across subjects (*columns*), showing the subject-specific coefficients for each component.

**Supplementary Table 1. Quality and performance measures for different component numbers**

| Rank | Sparseness Basis Matrix W | Sparseness Coefficient Matrix H | Silhouette width consensus matrix | Cophenetic correlation | Cophenetic dispersion | Residuals | Residual sum of squares (RSS), i.e. Frobenius norm | Explained variance | % additional variance explained over lower rank | % additional variance explained over lower rank in random data |
|---|---|---|---|---|---|---|---|---|---|---|
| 2 | 0.178 | 0.257 | 0.891 | 0.970 | 0.755 | 128.334 | 92.060 | 0.905 | n.a. | n.a. |
| 3 | 0.378 | 0.229 | 0.751 | 0.937 | 0.661 | 92.003 | 67.560 | 0.930 | 2.53% | 6.35% |
| 4 | 0.510 | 0.218 | 0.735 | 0.940 | 0.678 | 65.902 | 47.096 | 0.951 | 2.11% | 6.73% |
| 5 | 0.650 | 0.192 | 0.690 | 0.946 | 0.690 | 47.295 | 34.019 | 0.965 | 1.35% | 6.39% |
| **6** | **0.695** | **0.190** | **0.622** | **0.910** | **0.675** | **32.908** | **23.043** | **0.976** | **1.13%** | **5.93%** |
| 7 | 0.745 | 0.197 | 0.542 | 0.906 | 0.673 | 22.057 | 15.956 | 0.984 | 0.73% | 5.20% |
| 8 | 0.780 | 0.194 | 0.565 | 0.906 | 0.690 | 12.193 | 8.666 | 0.991 | 0.75% | 4.61% |
| 9 | 0.854 | 0.188 | 0.483 | 0.885 | 0.660 | 4.749 | 2.573 | 0.997 | 0.63% | 3.77% |
| 10 | 0.880 | 0.166 | 0.378 | 0.841 | 0.662 | 0.988 | 0.348 | 1.000 | 0.23% | 2.62% |
| 11 | 0.809 | 0.216 | 0.216 | 0.792 | 0.657 | 0.446 | 0.140 | 1.000 | 0.02% | 1.33% |

Table presents the metrics used to evaluate the NMF models using different ranks (*i.e.* components). The optimal number of components was selected based on: high sparseness of the W matrix (better interpretability), low cophenetic dispersion and high cophenetic correlation (clustering stability), low residual sum of squares (RSS) and residuals (better model fit), and a significant increase in explained variance compared to lower ranks and randomised data.

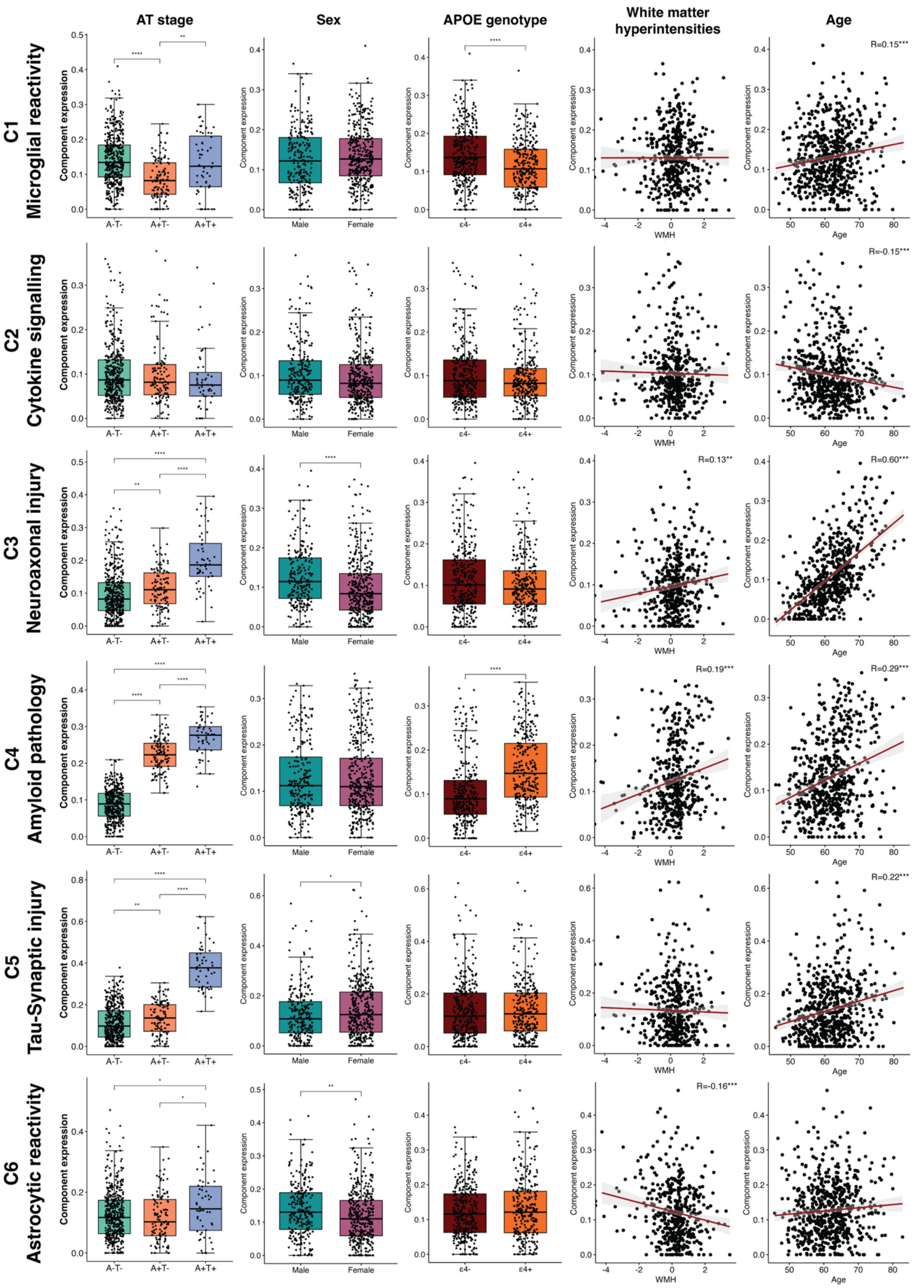


**Supplementary Figure 3. Association of biomarker components with AT stage, Age, Sex, *APOE ε4* carrier status, and WMH**

Boxplots show the relationship between component expression and AT stage, sex, and *APOE ε4* carrier status. Scatterplots showing the correlation of component expression with age and WMH (log-transformed). Significance levels: * $p < .05$, ** $p < .01$, *** $p < .001$.

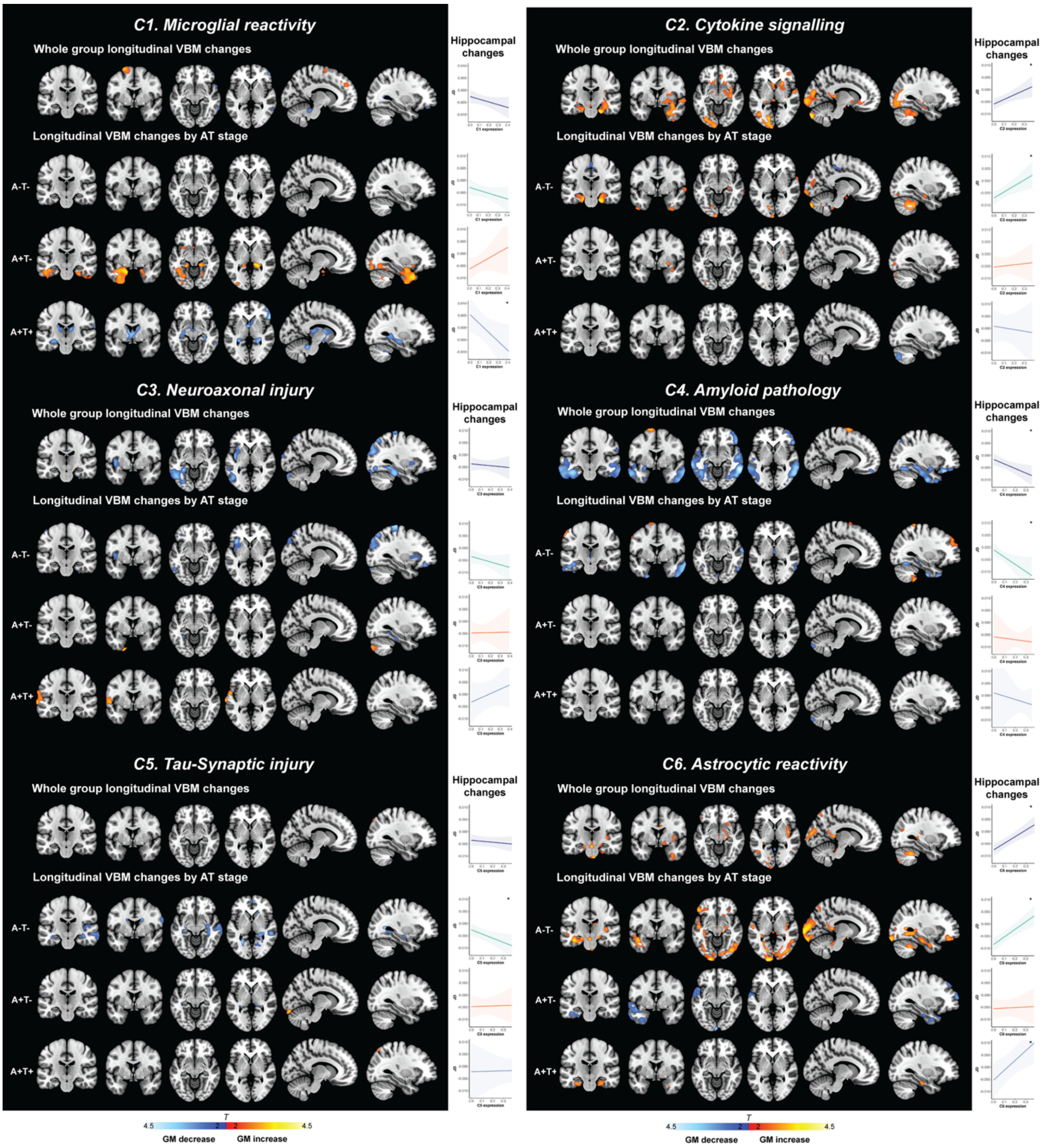


**Supplementary Figure 4. Voxel-wise associations of longitudinal grey matter volume changes with CSF biomarker components – mutually adjusted models.**

Overview of different biomarker components and their association with longitudinal grey-matter changes. Voxel-wise associations of longitudinal grey matter volume changes (*i.e.* Jacobian determinants [JD]) with biomarker components (C1-C6) in the entire sample and by AT stage, using a joint all model (all components entered simultaneously). Brain maps display regions where higher expression of components are associated with significant GM volume decreases (blue) or increases (red). Threshold: $p < 0.01$, $k > 100$). Complementary plots show the association between component (C1-C6) expression and longitudinal hippocampal change in the entire sample, and within AT subgroups. All analyses are adjusted for age, sex, *APOE ε4* carrier status, TIV, time interval between scans, and the other components. Significance level: * $p < .05$.

### Whole sample

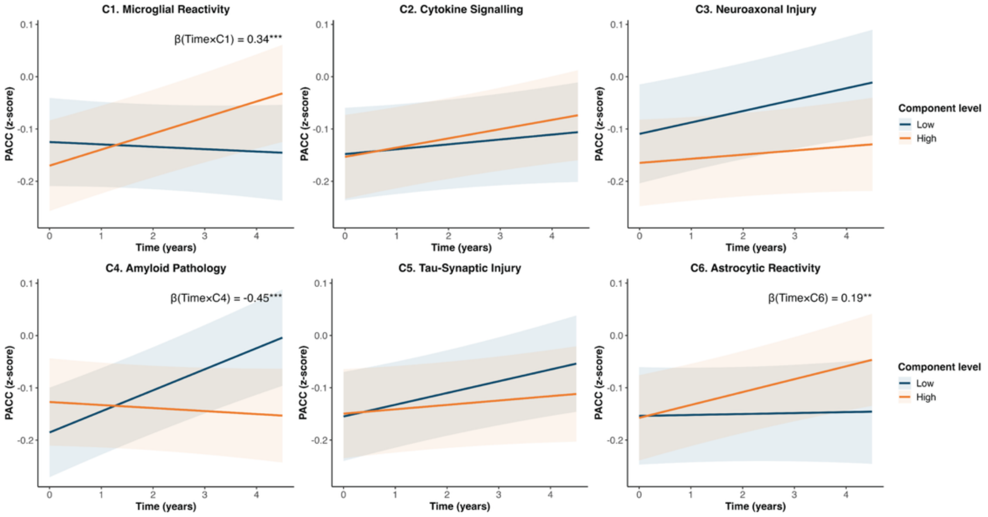


### Stratified by Aβ-status

#### AD continuum (Aβ+)

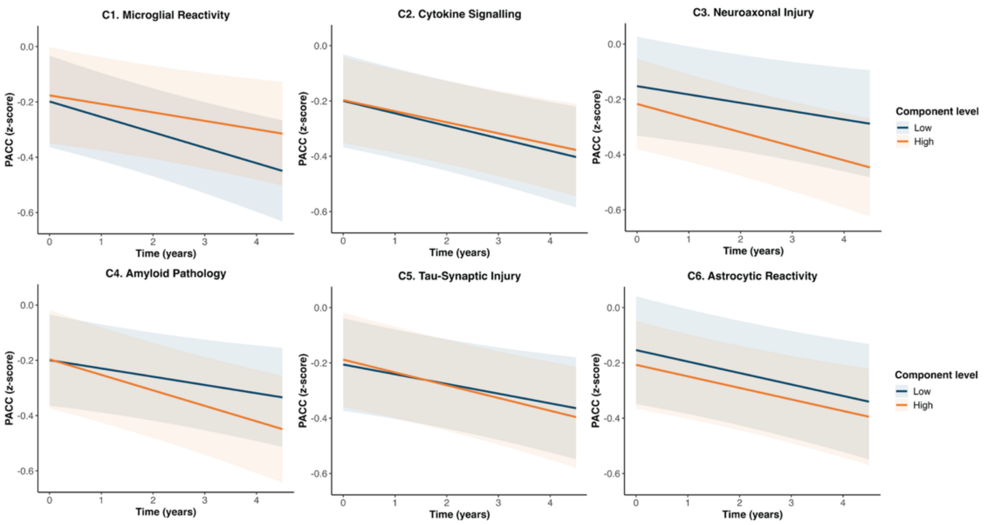


#### Controls (Aβ-)

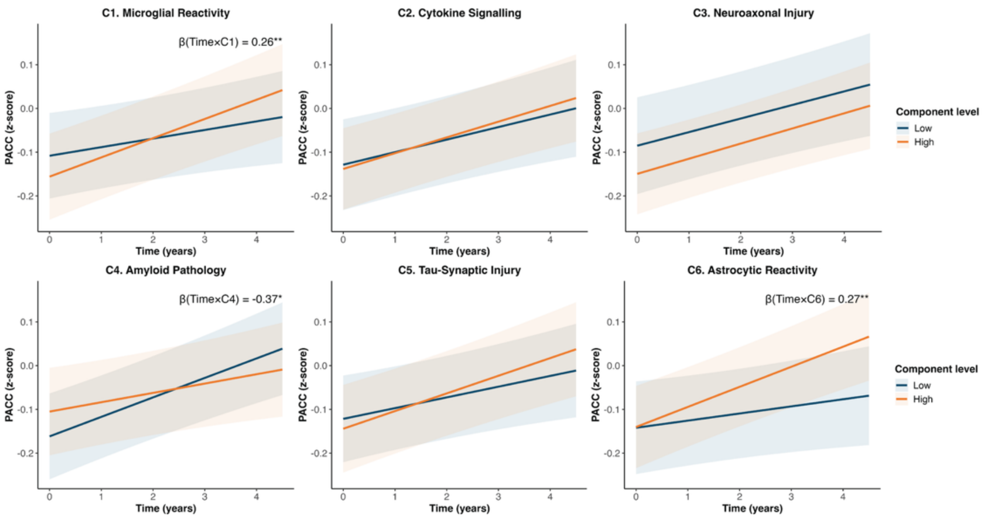

**Supplementary Figure 5. Association between biomarker component expression and longitudinal cognitive performance**

Cognitive trajectories were modelled using linear mixed-effect models with subject-specific random intercepts, across all participants (top) and, stratified by Aβ status (Bottom). The β-coefficients reflect significant Components x Time interactions on PACC scores, controlling for age, sex, education, and study centre. For visualisation, individuals were grouped into low (blue) or high (orange) component expression defined at $25^{th}$ and $75^{th}$ percentiles of the observed component distribution from population level predicted means and 95% confidence intervals. Individuals on the AD continuum defined by CSF Aβ42/40 ratios. * $p<.05$, ** $p<.01$, *** $p<.001$. PACC: Preclinical Alzheimer Cognitive Composite.

### Whole sample

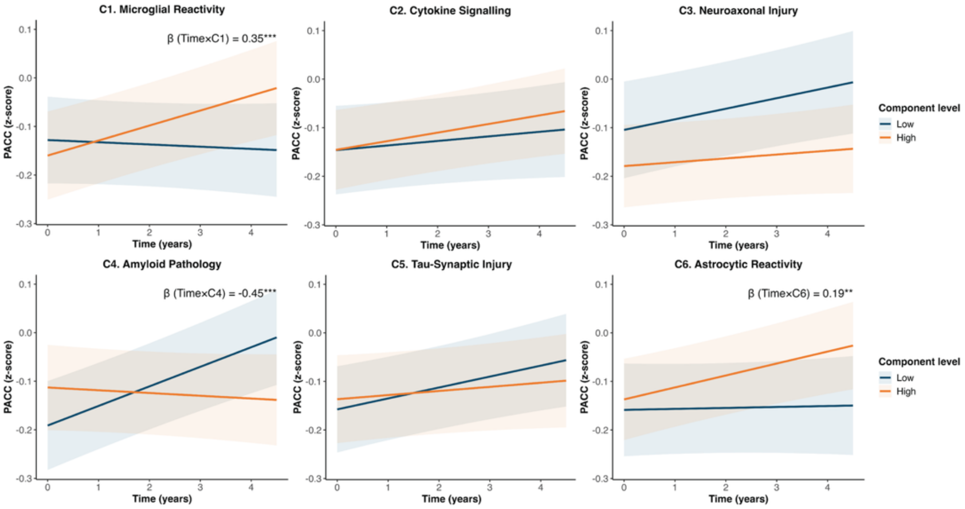


### Stratified by Aβ-status

#### AD continuum (Aβ+)

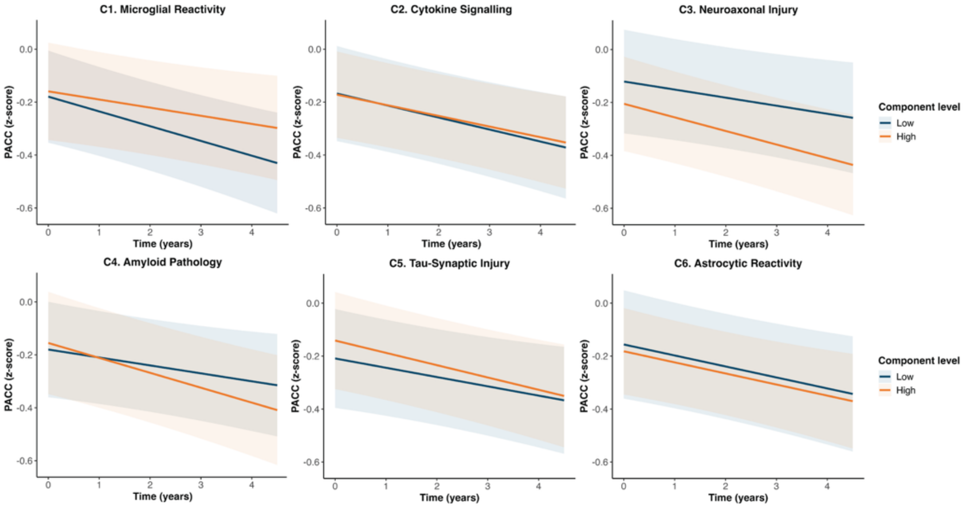


#### Controls (Aβ-)

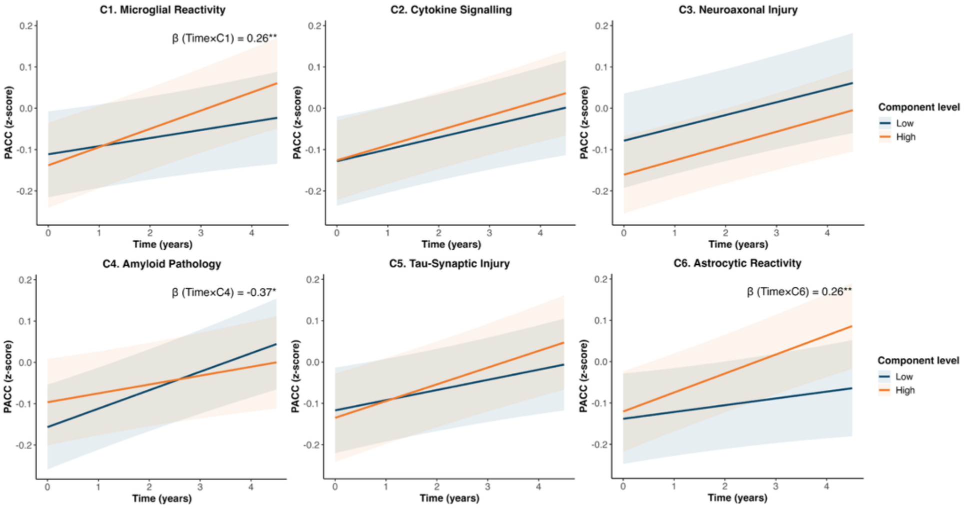

**Supplementary Figure 6. Association between biomarker component expression and longitudinal cognitive performance – mutually adjusted models.**

Cognitive trajectories were modelled using linear mixed-effect models with subject-specific random intercepts, across all participants (top) and, stratified by Aβ status (Bottom). The β-coefficients reflect significant Components x Time interactions on PACC scores, controlling for age, sex, education, study centre, and all other biomarker components. For visualisation, individuals were grouped into low (blue) or high (orange) component expression defined at $25^{th}$ and $75^{th}$ percentiles of the observed component distribution from population level predicted means and 95% confidence intervals. Individuals on the AD continuum defined by CSF Aβ42/40 ratios. * $p<.05$, ** $p<.01$, *** $p<.001$. PACC: Preclinical Alzheimer Cognitive Composite.
